# Phenome-wide association study of loci harboring *de novo* tandem repeat mutations in UK Biobank exomes

**DOI:** 10.1101/2022.01.26.22269821

**Authors:** Frank R Wendt, Gita A Pathak, Renato Polimanti

## Abstract

**Background:** Tandem repeats (TRs) are a major source of variation in the human genome under-investigated by large-scale genetic studies. When present in coding regions, TRs may have large effects on protein structure and function contributing to health and disease.

**Methods:** In a family-based design of 39 European ancestry trios from the UK Biobank (UKB), the GangSTR and MonSTR methods were used to identify *de novo* TRs in whole-exome sequences. TRs were annotated for association with gene expression and gene set enrichment. The loci harboring *de novo* TRs were investigated in a phenome-wide association study in up to 148,607 unrelated UKB participants of European descent. Linear and logistic regression included age, sex, sex×age, age^2^, sex×age^2^, and ten within-ancestry principal components as covariates. TR loci were fine-mapped to identify likely causal associations.

**Results:** There were 427 mutated TRs with a trend towards expansions versus contractions (*χ*^2^=5.46, df=1, *P*=0.019). These TRs were enriched for targets of the tumor suppressor microRNA-184 (21.1-fold, *P*=4.30×10^−5^). There were 123 TR-phenotype associations with posterior probabilities>0.95. These were related to body structure, cognition, and cardiovascular, metabolic, psychiatric, and respiratory outcomes. The most significant was between *NCOA6*-[GT]_N_ and “*ease of skin tanning*” (beta=0.069, se=0.003, *P*=1.51×10^−155^). There were several loci with large likely causal effects on tissue microstructure, including the association of *FAN1*-[TG]_N_ with carotid intima-media thickness (mean thickness: beta=5.22, se=1.08, *P*=1.22×10^−6^; maximum thickness: beta=6.44, se=1.32, *P*=1.12×10^−6^.

**Conclusions:** Combined with the TR *de novo* mutational background characterized herein, TR-phenotype associations contribute clear and testable hypotheses of dose-dependent TR implications linking genetic variation and protein structure with health and disease outcomes.

## INTRODUCTION

Tandem repetitive elements (TRs) are genomic loci consisting of consecutively repeated basepair motifs and represent one of the largest sources of genetic variation in humans [1]. TR motifs range from 1 to >20 basepairs and can be repeated dozens of times [2]. Variation in TR copy number has been associated with many diseases, including Huntington’s disease, which is characterized by over 100 copies of a CAG motif in *HTT* [2]. In combination, the need for large DNA sequencing datasets and the complexity of aligning repetitive DNA sequence reads to reference genomes [3-6] have contributed to the omission of TRs from large genome-wide studies of health and disease [7, 8]. Phenome-wide association studies of large TRs (i.e., TRs with >9 basepairs in the repeat unit) revealed likely causal effects on height, hair morphology, and several biomarkers of human health [8]. Most notably, these TRs have substantially larger, and independent, effects on phenotype relative to nearby SNPs detected by genome-wide association studies. To date, small TRs between 1-9 basepairs have not been investigated for phenotypic consequences in large cohorts.

*De novo* mutations are a class of genetic variation where offspring harbor an allele absent in either parent. Single nucleotide *de novo* mutations are often rare, deleterious, and may disrupt gene function contributing to many genetic disorders [9]. With respect to complex traits, the cumulative burden of *de novo* mutations may play a large additive role in disease etiology [10]. Due to their repetitive nature, TRs have much higher mutation rates than single-nucleotide variants making *de novo* mutational events more common for this class of genomic variation [11]. *De novo* TRs have been associated with complex traits, like autism spectrum disorder, in family trios [12]. Often studied in the context of a specific condition, it is unclear how pervasive *de novo* TR mutations are across the genome and how they affect human health and disease.

In this study, we applied a two-staged analytic approach to UK Biobank (UKB) data to (i) identify *de novo* TR mutations using a family-based design and (ii) characterize the effect of variation in these TRs on 1,844 human traits using a population-based design. We report 426 TR associations with body structure, cognition, and cardiovascular, metabolic, psychiatric, and respiratory outcomes. Fine-mapping these loci identified 41 TRs with a high probability of large causal effects on structural features of the carotid artery and thalamus radiation, highlighting the critical role of TR variation on health and disease outcomes.

## SUBJECTS AND METHODS

### UK Biobank

The UKB is a large population-based cohort of >500,000 participants. UKB assesses a wide range of factors in including physical health, anthropometric measurements, circulating biomarkers, and sociodemographic characteristics [13]. This research has been conducted in the scope of UKB application reference number 58146. Individual-level data are available to bona fide researchers through approved access. Ancestry in the UKB was assigned using a random forest classifier based on features predictive of ancestry in a combined 1000 Genomes Project plus Human Genome Diversity Panel reference [14]. This procedure resulted in 174,371 European, 2,885 African, 1,220 East Asian, 4,107 Centra/South Asian, 692 Middle Eastern, 442 Admixed American participants with whole-exome sequencing (WES) data.

### Family trios

To determine relatedness among participants with WES data, we performed identify-by-descent (IBD) analysis per ancestry group. Linkage disequilibrium (LD) independent SNPs with minor allele frequency > 5% were clumped in Plink v1.9 [15] using 200-kb window size per 100 variants and a pairwise-*r*^*2*^ of 0.2. Pairwise IBD was performed in Plink using the --genome flag. With all pairs of individuals with pi_hat > 0.2, we used age, sex, and IBD metrics to determine parent-offspring relationships. The following cutoffs were applied to IBD metrics: 0 ≤ Z0 ≤ 0.05, 0.75 ≤ Z1 ≤ 1, and 0 ≤ Z2 ≤ 0.05 [16].

### De novo variant calling

UKB CRAM files containing WES reads aligned to the hg38 reference genome were converted to binary alignment map (BAM) files using cramtools v3.0 (February 2021) [17]. Genotyping of autosomal tandem repeats from short reads was performed with GangSTR v2.5.0 [18] using sorted and indexed BAMs. Each family trio was processed with a separate GangSTR job resulting in per-family variant call files (VCF). Per-family VCFs were subjected to quality control using dumpSTR [19] to remove TRs with (i) read depths <20X, (ii) reads that only span or flank the genotyped region, and (iii) maximum likelihood genotypes outside the 95% confidence interval reported by GangSTR.

MonSTR v1.1.0 [12] was used to identify *de novo* TRs per family. MonSTR evaluates the joint likelihood of genotypes in a parent-offspring trio to estimate a posterior probability of mutation at each TR in the offspring. MonSTR was run per family with non-default parameters as performed previously including the following flags: --max-num-alleles 100, --gangstr, --min-total- encl 10, --posterior-threshold 0.5, and --default-prior −3 [12]. In combination, these flags remove error-prone TRs, indicate to use GangSTR-output likelihoods, apply a constant prior of per-locus mutation rate set to 10^−3^, require *de novo* mutations to be supported by at least three enclosing reads, require a minimum of 10 enclosing reads per sample in the family, and label calls with posterior probability ≥ 0.5 as mutations.

### Gene annotation

TR mutations were positionally mapped to genes using the hg38 reference genome. To identify LD-independent genomic regions, we retained only one TR-containing gene in a 200-kb window. When TR-containing genes were closer than 200-kb [20], we retained the gene with the larger number of *de novo* mutations observed. Hypergeometric tests were used to determine gene set enrichments of gene ontologies from the Molecular Signatures Database. Multiple testing (FDR 5%) correction as applied per MSigDB category (N=17 categories) [21]. The number of gene-sets per category ranged from 50 (Hallmark gene sets) to 5,219 (Immunologic). The same gene-sets were then tested with a second method, ShinyGo [22], which was used as a complementary method because it reports the magnitude of the fold change associated with each gene set. ShinyGo reports *P*-values that have been adjusted using a false discovery rate of 5% considering all gene sets tested.

### Expression associated TRs

Gene expression associated TRs (eSTR) were identified using data from Fotsing, et al. [20]. Briefly, average TR repeat length, called from whole genome sequences using HipSTR [23], was associated with gene expression in 17 tissues from the Genotype-Tissue Expression Project (N=652 unrelated individuals; 86% European ancestry). All STR-gene expression relationships were adjusted for age, sex, and the top 10 within-ancestry principal components.

### Phenome-wide association (PheWAS)

PheWAS was performed in up to 148,607 unrelated UKB participants of European ancestry based on the random forest classifier applied in the Pan-ancestry UK Biobank project [14]. Linear and logistic regression models were applied to continuous and binary outcomes, respectively, with the requirement for at least 1,500 cases for binary traits (∼1% of the total sample size). TR loci with *de novo* mutations were called from WES data using GangSTR, as described above in *De novo variant calling*. PheWAS was performed for all loci harboring *de novo* TRs requiring a mean read depth >20X and missingness <5% across all unrelated European ancestry participants.

After trait, individual, and TR quality control steps, PheWAS was performed for 416 TRs across 1,844 phenotypes. To associate multi-allelic TRs with traits of interest, we converted TR genotypes to a TR length sum which captures locus-level burden. The length sum per TR was calculated by adding the number of repeat units in a genotype. Prior to regression, the TR length sum per locus was normalized/residualized using age, sex, sex×age, age^2^, sex×age^2^, and the top 10 within-ancestry principal components as covariates. The generalized linear model function in R was used to regress normalized/residualized TR length burden on each phenotype. Multiple testing correction was applied using the false discovery method (false discovery rate (FDR)<5%) to accommodate the correlation between traits and between TRs.

### Fine-mapping TR-SNP signals

To determine if TR-phenotype association signals represented likely causal relationships between TR variation, rather than tagging nearby SNP signals, we fine-mapped each TR-containing locus [8]. We computed the linear regression between TR length sum and all SNPs and insertion/deletion (indel) polymorphisms within 500-kb of the TR start site. Bayesian fine-mapping was performed with FINEMAP v1.3.1 [24] with the optional flags: --corr-config 0.999 -- sss --n-causal-snps 5. These settings estimate the likelihood of causality for the TR accounting for LD with other genetic variants in the region. SNP and indel effect sizes per trait were estimated by linear or logistic regression in the same sample from which TR effect sizes were estimated. Each SNP/indel effect size estimate included age, sex, sex×age, age^2^, sex×age^2^, and the top 10 within-ancestry principal components as covariates.

## RESULTS

### Family trios and de novo mutations

After IBD analysis and quality control, 40 trios were identified (39 EUR and 1 AFR). There were 17 male and 22 female offspring (*χ*^2^=0.119, df=1, *P*=0.423) with no difference in age between the sexes (male mean=42.60±1.94, female mean=43.05±1.79, *t*=-9.23, df=35.30, *P*=0.362). Due to sample size limitations in diverse ancestry trios, the genetic analyses focus only on the 39 trios of EUR descent (Table S1).

Five families had no detectable *de novo* TR mutations after applying the quality thresholds for this study. Across 34 EUR families, there were 1,031 *de novo* TR mutations (mean number of TRs per family=31.2±18.5; Table S2). These 1,031 mutations reflect 427 loci, 211 of which were mutated in >1 trio (Figure 1). There was a significant trend towards TR expansions *versus* contractions (*χ*^2^=5.46, df=1, *P*=0.019). The mean expansion size was 1.41±0.98 repeat units (maximum mutation was 6 repeat units at *HTT*-[CAG]_N_) and the mean contraction size was - 1.34±0.69 repeat units (maximum mutation was −12 repeat units at *DST*-[CA]_N_). The most frequently mutated TR was *NOTCH4*-[AGC]_N_, observed in 11/39 trios and mutated between 1 and 3 repeat units per trio.

**Figure 1.**
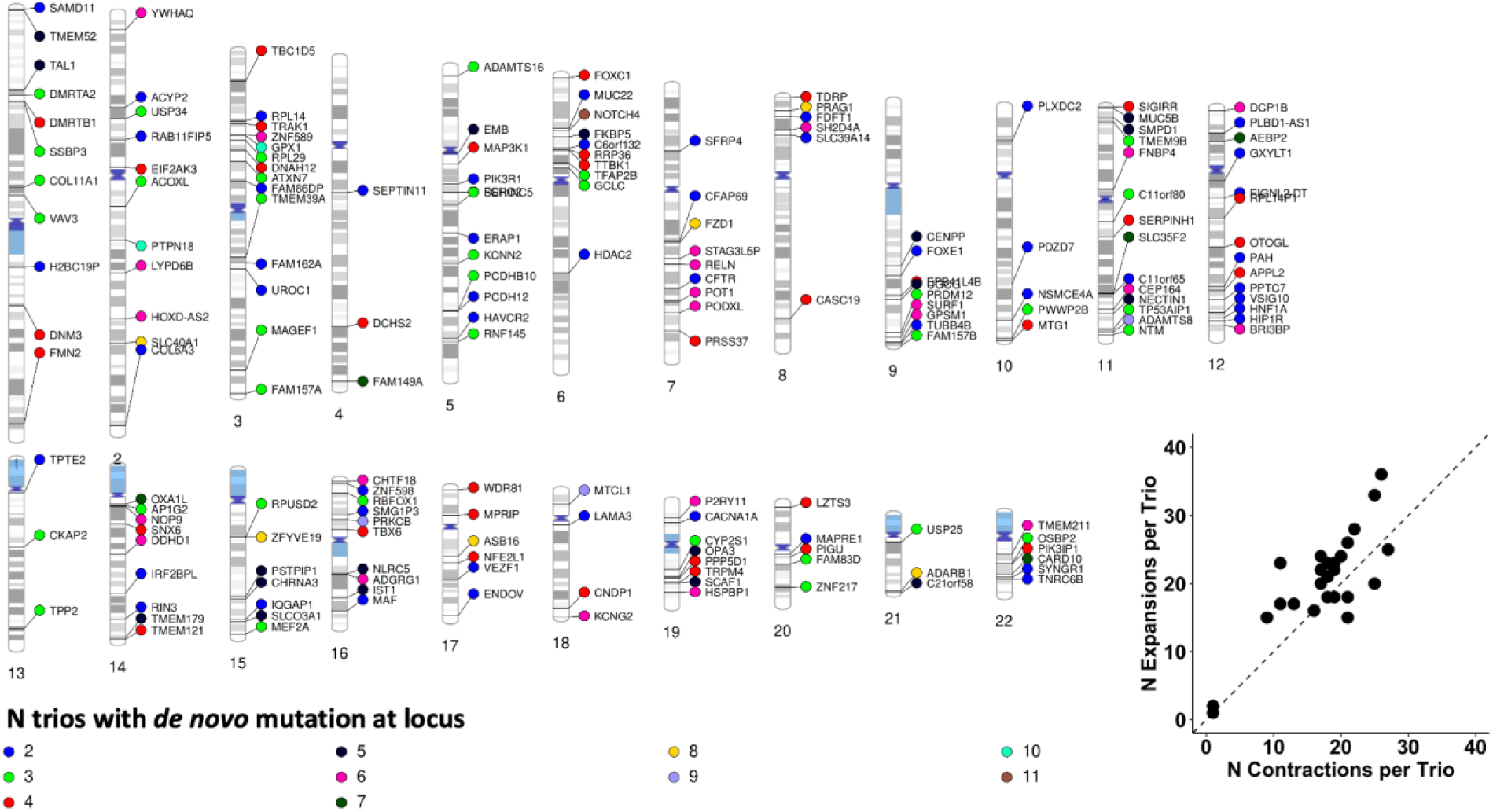
Chromosomal localization of *de novo* TR mutations observed in at least two trios. Each data point is a single TR mutation color coded to designate the number of UKB European ancestry trios in which that mutation was observed. Each data point is annotated with the gene containing the TR locus. Loci are positioned relative to the hg38 reference genome. The bottom right inset shows the slight bias in repeat expansions relative to repeat contractions per trio (N=39). The diagonal dashed line indicates a 1:1 ratio of TR expansions:contractions.

### Locus annotation

We applied two methods to annotate the TR loci with *de novo* mutations. First, we tested whether each TR was associated with the expression of the positionally mapped TR-containing gene. A set of 352 approximately LD-independent TRs was selected by selecting a single TR in a 200-kb window, prioritizing the locus with the larger number of *de novo* mutations in this dataset. Using these 352 genes, we performed hypergeometric tests for gene set enrichment in FUMA and ShinyGo. After multiple testing correction (FDR 5% performed per gene set category), there were eight gene sets significantly enriched in FUMA and ShinyGO (Table S3). The largest enrichment was related to targets of MIR184 (21.1-fold enrichment, FUMA *P*=4.30×10^−5^, SinyGO FDR-adjusted *P*=0.01). The remaining enrichments include growth hormone pathway, cell morphogenesis involved in differentiation (GO:0000904), targets of microRNAs 515-1, 515-2, and 519E, targets of transcription factors SP1 and SOX9, genes downregulated in regulatory T-cells relative to conventional T-cells (GSE13306), and genes involved in epithelial-mesenchymal transition (MSigDB M5930).

Next, expression-associated TRs were analyzed with respect to 17 tissues in the GTEx repository [20]. From the same set of 352 approximately LD-independent TRs, there were 259 significant TR-gene expression associations across 17 tissues (Figure 2). Among them, *CKAP2*-[GCGGTG]_N_ was associated with expression across 17 tissues (Z-score range 3.26 in transformed fibroblasts to 11.77 in tibial artery).

**Figure 2.**
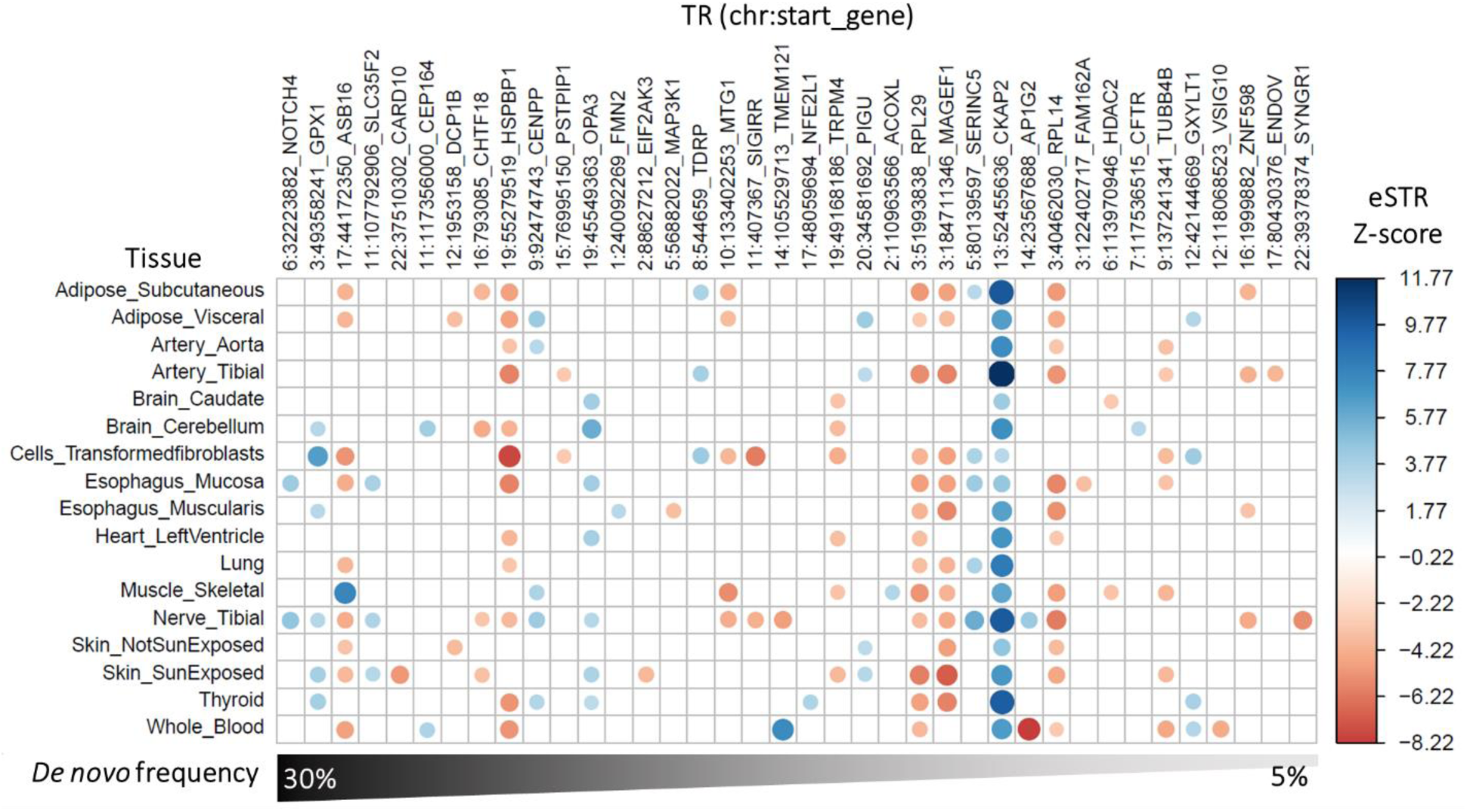
Association between tandem repeats (TR) and gene expression across 17 tissues from GTEx [20]. Each TR is named according to its start position with respect to the hg38 reference genome and the gene containing the TR whose expression is influenced by variation at the TR. The heatmap is restricted to the TRs harboring *de novo* mutations in more than one trio (∼5% frequency). All TR-gene-tissue association statistics are provided in Table S4.

### PheWAS of TRs harboring de novo mutations

To characterize the phenome-wide effects of each TR exhibiting a *de novo* mutation, we associated TR length sum with 1,844 phenotypes in up to 148,607 European ancestry participants. After removing TRs with low mean read depth (4 TRs) and high missingness rates (8 TRs), PheWAS was performed for 415 TRs. After FDR multiple testing correction (*P*<2.75×10^−5^), there were 426 TR-phenotype associations, representing 97 TRs (Figure 3 and Table S5). The most significant and likely causal association was between *NCOA6*-[GT]_N_ and “*ease of skin tanning*” (UKB Field ID 1727) such that individuals who burn rather than tan tend to have longer alleles at this locus (beta=0.069, se=0.003, *P*=1.51×10^−155^, posterior probability=1). The maximum Cohen’s *d* comparing effect sizes across “*ease of skin tanning*” categories was 0.212 (“*Get very tan*” versus “*Never tan, only burn*” P=9.21×10^−308^; Figure 4A and Table S6).

**Figure 3.**
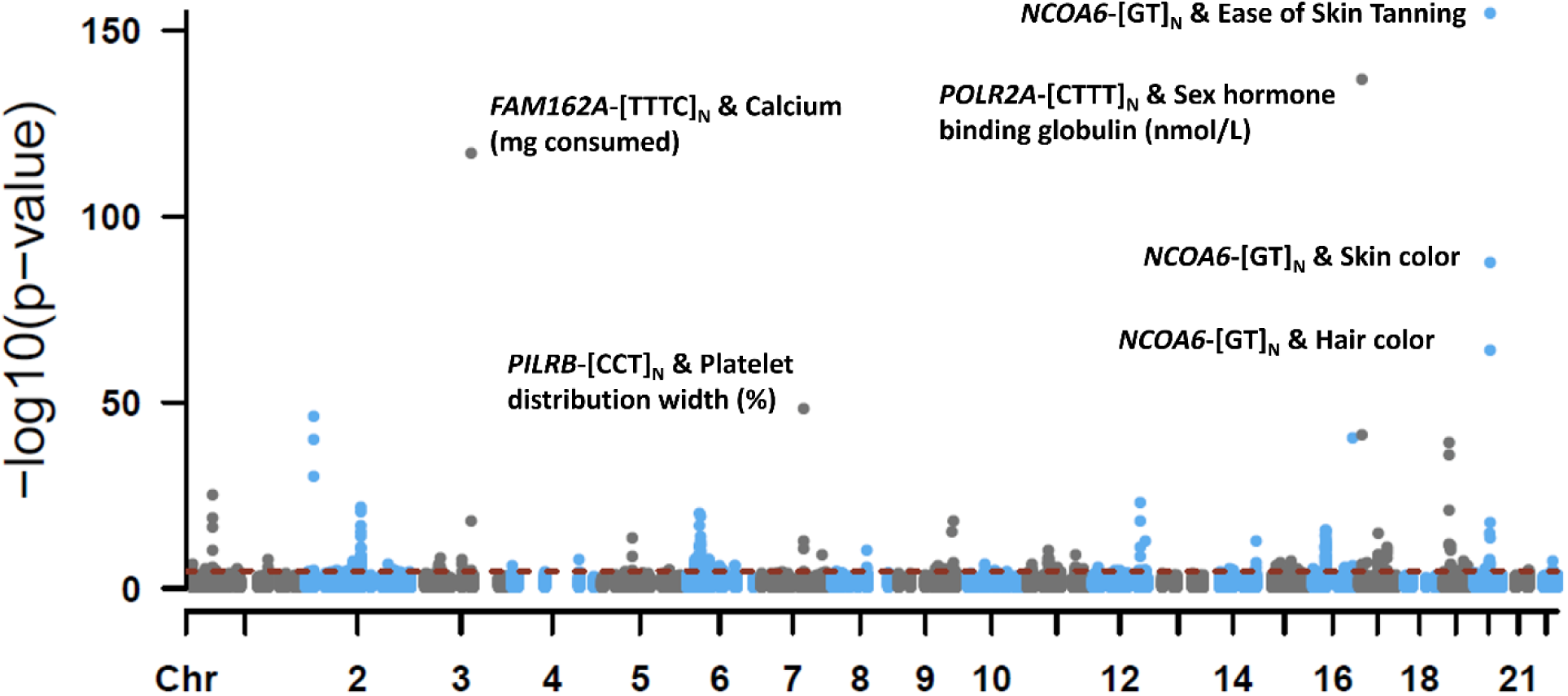
Manhattan plot of phenome-wide association results for 415 TRs and 1,844 phenotypes in European ancestry participants of the UK Biobank. The dashed red line indicates the P-value threshold after multiple testing correction (FDR<5%; *P*=2.75×10^−5^). Select TR-phenotype associations are labeled. All summary statistics are provided in Table S5.

**Figure 4.**
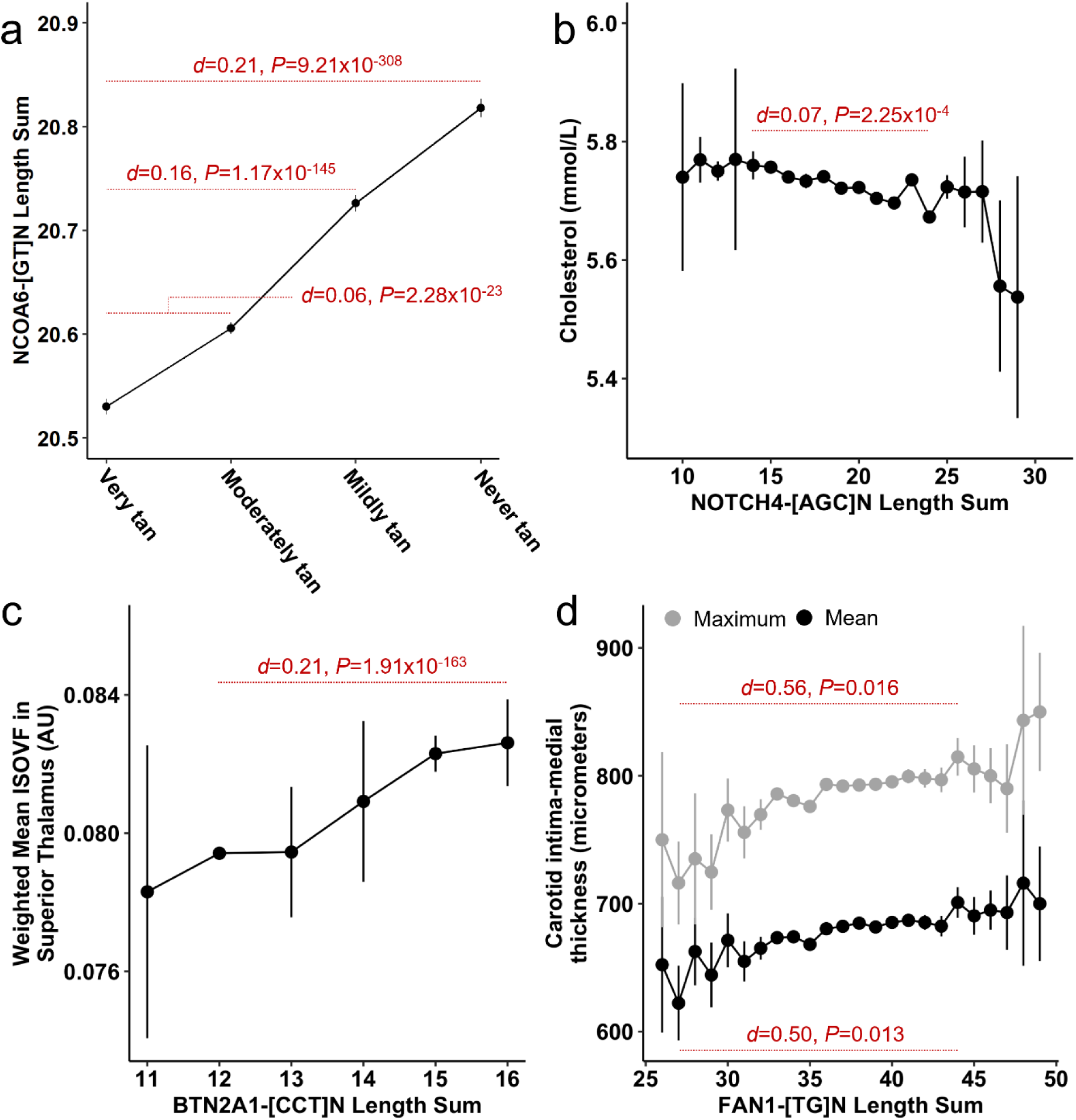
Effects of TR length burden on various phenotypes in UK Biobank participants of European ancestry. Each data point represents the mean (± standard error) phenotype measurement at the indicated TR length. Red text denotes several small to medium effect size differences (Cohen’s *d* and *P*-value) between phenotype estimates.

For the 27 TRs with significant (FDR<5%) associations across multiple trait domains, hypergeometric tests were applied to identify enriched domains (Table S7). Twelve TRs were enriched for associations with hematological measures (mean fold-enrichment 57.9±55.8), the most significant of which was *NOTCH4*-[AGC]_N_ (64.5-fold enrichment of hematology associations, *P*=3.28×10^−16^). The largest likely causal effect size observed for this locus was with respect to “*total cholesterol*” (UKB Field ID 30690) such that individuals with longer TR length have lower cholesterol concentration (beta=-0.014, se=0.003, *P*=5.91×10^−6^, posterior probability=1). The maximum Cohen’s *d* comparing effect sizes across *NOTCH4*-[AGC]_N_ length sum was 0.074 (*NOTCH4*-[AGC]_15_ versus *NOTCH4*-[AGC]_24_ *P*=2.25×10^−4^; Figure 4B and Table S6).

### Notable likely causal TR-phenotype associations

After fine-mapping, there were 123 TR-phenotype associations involving 41 TRs with evidence that variation in the TR (posterior probability > 0.95), rather than a nearby SNP or indel, drove the genotype-phenotype association (Table S8). Though not enriched among the *de novo* variants detected, there were notable relatively large effects and likely causal TRs associated with structural components of the white matter microstructure and peripheral vasculature. Variation in *BTN2A1*-[CCT]_N_ was associated with increased isotropic volume fraction (ISOVF) in the right superior thalamic radiation (beta=7.33×10^−4^, se=1.41×10^−4^, P=2.19×10^−7^; posterior probability=0.989). The largest difference was observed between *BTN2A1*-[CCT]_12_ and *BTN2A1*-[CCT]_16_ (Cohen’s *d*=0.209; *P*=1.91×10^−163^; Figure 4C). Length variation at *FAN1*-[TG]_N_ was associated with the mean (beta=5.22, se=1.08, *P*=1.22×10^−6^; posterior probability=0.979) and maximum (beta=6.44, se=1.32, *P*=1.12×10^−6^; posterior probability=0.985) carotid intima-media thickness (IMT). For mean (Cohen’s *d*=0.50, *P*=0.013) and maximum (Cohen’s *d*=0.56, *P*=0.016) carotid intima-media thickness, the largest effect size was observed between *FAN1*-[TG]_27_ and *FAN1*-[TG]_44_ (Figure 4D).

## DISCUSSION

TRs are a major source of genetic variation in humans that are under-investigated in common complex traits but have great potential to inform the causal biology of disease states when localized to regulatory and coding regions of the genome. Due to their repetitive nature, TRs have relatively high mutation rates that enrich this class of variation for *de novo* mutations. Leveraging a two-staged family-based and population-based design, this study identified 427 loci harboring *de novo* TR mutations. There were 426 TR-phenotype associations across several health and disease domains with significant enrichment of loci associated with biomarkers such as cholesterol and alkaline phosphatase concentrations. Notably, there were 124 TR-phenotype associations where the TR, not surrounding SNPs or indels, was likely causal for trait variation thereby highlighting a large contribution to the genetic etiology of health and disease that is independent of signals identified by GWAS.

In a sample of European ancestry family trios from the UKB, we identified 427 loci harboring *de novo* TR mutations. The most frequently mutated TR mapped to *NOTCH4. NOTCH4* encodes neurogenic locus notch homolog 4 and is almost exclusively expressed in the vasculature where changes in gene expression produce less vasculature branching and reduced vessel integrity. NOTCH4 has been the subject of many studies of schizophrenia. However, the TR locus identified here is contested as a causal variant in the gene [25]. *NOTCH4*-[AGC]_N_ variation was enriched for association with biomarkers and metabolic health, consistent with its localization to the vasculature. The strongest result identified longer copies of the *NOTCH4*-[AGC]_N_ TR associated with lower total cholesterol concentration. The polyleucine stretch encoded by this TR makes up a portion of the NOTCH4 extracellular domain and therefore may contribute to abnormal astrocyte differentiation, angiogenesis, and coronary vessel development [26].

As a whole, the collection of genes harboring mutated TRs was enriched for targets of the transcriptionally relevant micro-RNAs 184 and 5155P-519E and transcription factors Sp1 and SOX9. Micro-RNA 184 is a tumor suppressor implicated in numerous cancers as it plays a critical role in cell differentiation and fate across tissues [27-29]. SOX9 is often overexpressed in various solid tumors and its upregulation contributes [30] to the mutability of cancerous cells while Sp1 is a ubiquitously expressed transcription factor with binding sites in the promoter regions of many cell cycle regulatory proteins [31]. In combination, these observations support the role of genome instability in the origin of *de novo* variation while characterizing a background of *de novo* TR mutations in the UKB cohort.

The most significant PheWAS finding related *NCOA6*-[GT]_N_ and “*ease of skin tanning*.” This TR was enriched for association with dermatological phenotypes and was determined likely causal for ease of skin tanning, hair color, and skin color, with effects independent of all surrounding SNPs and indels. NCOA6 gene expression has been previously associated with outwardly visible characteristics [32] and variation in this locus is part of predictive algorithms for freckling in Europeans [33]. Here we describe the likely causal effect of a TR that may hold additional predictive properties that explain the poor genetic prediction of intermediate phenotypes. For example, SNP-based skin color predictions poorly perform with pale and intermediate skin tones but predict darker skin tones relatively well [34].

Fine-mapping TR loci revealed approximately 20% of the TR-phenotype associations detected were likely causal such that nearby SNPs and indels were less likely to explain phenotypic variation than the TR itself. Two TRs had relatively large effects on biological features relevant for human health and disease: *BTN2A1*-[CCT]_N_ associated with isotropic volume fraction in the thalamus and *FAN1*-[TG]_N_ associated with the mean and maximum carotid intima-media thickness. BTN2A1 is a subtype of butyrophilin, an immunoglobulin family associated with chronic kidney disease, ulcerative colitis, and rheumatoid arthritis [35]. Here we demonstrate that longer repeat copy number in *BTN2A1*-[CCT]_N_ associated with greater ISOVF in the superior thalamus, a region of the brain also implicated in chronic kidney disease [36], ulcerative colitis [37], and common psychiatric conditions like depression [38]. These pleiotropic effects link comorbid conditions to clear biological mechanisms (i.e., longer repeat lengths correspond to larger thalamus ISOVF).

FAN1 encodes a DNA repair nuclease and has been previously implicated in several TR-associated health outcomes [39]. The most notable is the polyglutamine expansions underlying Huntington’s disease and spinocerebellar ataxia type 1 [40]. Though itself a nuclease, FAN1 may modulate repeat instability through a nuclease-independent mechanism related to its own genetic variation but other data suggest certain aspects of nuclease activity contribute to repeat instability [41]. The *FAN1*-[TG]_N_ TR was associated with a likely causal effect on carotid IMT independent of common SNPs that empirically influence FAN1 activity (e.g., rs3512 and rs35811129) [39]. The large effect at this locus corresponds to a 78.7-micrometer difference in mean and 98.6-micrometer difference in maximum carotid IMT. Carotid IMT above 880-micrometers has been ascribed atherosclerosis diagnostic sensitivity and specificity up to 88% and 90%, respectively [42], though these metrics range with demographics of the sample. Taken together, genotyping the *FAN1*-[TG]_N_ TR may have the potential to increase the diagnostic accuracy of atherosclerosis given carotid IMT data [43].

This study has some limitations to consider. First, TRs are abundant throughout the genome and similar to SNP signals detected by GWAS, non-coding TRs may play regulatory roles through trans-QTL effects that cannot be captured by exome-focused studies of the genome. Furthermore, the complex traits assessed herein are highly polygenic and although several TRs identified in this study have relatively high effects, they likely act in aggregate rather than isolation. Future studies are required to model the polygenic versus monogenic burden of TRs, SNPs, and the combination of TRs and SNPs. Second, the *de novo* mutation background characterized in this study is likely underestimated due to the relatively small number of trios and the focus on European ancestry participants. Future studies will leverage the full extent of family data and ancestry diversity in the UKB to quantify the diversity of *de novo* TR mutations and their cross-ancestry heterogeneity.

## CONCLUSIONS

This study provides an atlas of the phenotypic spectrum associated with loci harboring *de novo* TR mutations in the UKB. Independent of the surrounding biallelic genetic variation, the TRs detected here have relatively large effects on phenotypic outcomes and generally localize to regions that translate to amino acid stretches in proteins. Therefore, these associations serve as testable hypotheses regarding the size and copy number effects of TR loci on health and disease with clear dose-dependent implications. These findings will contribute to the mechanisms linking genetic variation, protein structure, and health and disease outcomes.

## Data Availability

The data supporting the conclusions of this article are included within the article and its additional files.

## ABBREVIATIONS

*HTT*: Huntingtin encoding gene
TR: tandem repeat
DNA: deoxyribonucleic acid
SNP: single nucleotide polymorphism
UKB: UK Biobank
WES: whole exome sequencing
IBD: identity by descent
kb: kilobase
BAM: binary alignment map
VCF: variant call format
LD: linkage disequilibrium
FDR: false discovery rate
MSigDB: Molecular Signatures Database
eSTR: expression associated short tandem repeat
PheWAS: phenome-wide association study
*DST*: dystonin encoding gene
*NOTCH4*: notch receptor 4 encoding gene
FUMA: Functional Mapping and Annotation
MIR184: microRNA-184
GO: gene ontology
*SOX9*: SRY-Box Transcription Factor 9 encoding gene
*Sp1*: specificity protein 1 encoding gene
GTEx: Genotype-Tissue Expression
*CKAP2*: cytoskeleton-associated protein 2 encoding gene
*NCOA6*: nuclear receptor coactivator 6 encoding gene
*BTN2A1*: butyrophilin subfamily 2 member A1 encoding gene
*FAN1*: FANCD2 And FANCI Associated Nuclease 1 encoding gene
IMT: intima-media thickness
ISOVF: isotropic volume fraction

## DECLARATIONS

This research was conducted using the UK Biobank Resource (application reference no. 58146). The authors thank the research participants and employees of the UK Biobank for making this work possible. This study was supported by the National Institutes of Health (R21 DC018098, R33 DA047527, and F32 MH122058).

## COMPETING INTERESTS

There are no competing interests to report.

## AVAILABILITY OF DATA AND MATERIALS

The datasets supporting the conclusions of this article are included within the article and its additional files.

## SUPPLEMENTARY TABLES

Table S1. European family trios. Seven trios had no detectable *de novo* TR mutations based on the quality control thresholds applied in this study.

Table S2. All *de novo* TR mutations detected across 39 UKB family trios of European descent.

Table S3. Gene-sets enriched for loci harboring de novo TR mutations using ShinyGO and FUMA. Gene sets highlighted in yellow are significant after multiple testing correction using both methods.

Table S4. Expression-associated TRs across 17 tissue types from GTEx. Empty cells indicate that a TR was not present in the TR-expression study performed by Fotsing, et al. [20].

Table S5. All significant associations between TRs and UK Biobank phenotypes. Each row describes a single TR-phenotype association. The UKB Field ID, full trait description, and trait domain are provided.

Table S6. Comparison of TR length sum effects for select TR-phenotype associations. Cohen’s *d* is reported for each outcome comparing each pair of TR lengths with at least 7 observations. Associated *P*-values were calculated using Z-tests.

Table S7. Per-locus enrichment of associated trait domains using hypergeometric tests. Highlighted cells indicate significant enrichment after multiple testing correction (FDR<5%).

Table S8. Significant fine-mapping results for TR-phenotype associations from Table S5.

## Notes

### Competing Interest Statement

The authors have declared no competing interest.

### Author Declarations

This research was conducted using the UK Biobank Resource (application reference no. 58146). UKB data can be accessed by bona fide researchers through the UKB data access portal.

